# A typology of physician input approaches to using AI chatbots for clinical decision-making: a mixed methods study

**DOI:** 10.1101/2025.07.23.25332002

**Authors:** Rachel Siden, Hannah Kerman, Robert J. Gallo, Joséphine A. Cool, Jason Hom, Ethan Goh, Neera Ahuja, Paul Heidenreich, Lisa Shieh, Daniel Yang, Jonathan H Chen, Adam Rodman, Laura M Holdsworth

## Abstract

**Background:** Large language model (LLM) chatbots demonstrate high degrees of accuracy, yet recent studies found that physicians using these same chatbots may score no better to worse on clinical reasoning tests compared to the chatbot performing alone with researcher-curated prompts. It is unknown how physicians approach inputting information into chatbots.

**Objective:** This study aimed to identify how physicians interacted with LLM chatbots on clinical reasoning tasks to create a typology of input approaches, exploring whether input approach type was associated with improved clinical reasoning performance.

**Methods:** We carried out a mixed methods study in three steps. First, we conducted semi-structured interviews with U.S. physicians on experiences using an LLM chatbot and analyzed transcripts using the Framework Method to develop a typology based on input patterns. Next, we analyzed the chat logs of physicians who used a chatbot while solving clinical cases, categorizing each case to an input approach type. Lastly, we used a linear mixed-effects model to compare each input approach type with performance on the clinical cases.

**Results:** We identified four input approach types based on patterns of “content amount”: copy-paster (entire case), selective copy-paster (pieces of a case), summarizer (user-generated case summary), and searcher (short queries). Copy-pasting and searching were utilized most. No single type was associated with scoring higher on clinical cases.

**Discussion:** This study adds to our understanding of how physicians approach using chatbots and identifies ways in which physicians intuitively interact with chatbots.

**Conclusions:** Purposeful training and support is needed to help physicians effectively use emerging AI technologies and realize their potential for supporting safe and effective medical decision-making in practice.

## BACKGROUND

Large language model (LLM) chatbots (such as ChatGPT, Perplexity, or Claude) that users can interact with conversationally have proliferated in recent years,^1^ including in clinical contexts, and are increasingly being used for generating differential diagnoses.^2,3^ The most recent generation of LLM chatbots have demonstrated even greater abilities in addressing complex clinical cases with a high degree of accuracy in controlled settings with researcher-created prompts.^4,5^

The inputs to LLM chatbots can significantly influence the accuracy of outputs produced.^6,7^ Prompt engineering, the practice of optimizing LLM chatbot outputs with structured inputs, has emerged as an important field of study.^8^ However, prompt engineering best practices are not always intuitive to lay or untrained users, who may employ a variety of different input styles.^9,10^ When evaluating LLM chatbot outputs for accuracy and applicability in clinical settings, it will be important to consider not just how LLM chatbots perform under ideal conditions, but also how well they perform with everyday human users. In previous work evaluating the accuracy of chatbot responses to clinical cases, researchers found that a LLM chatbot alone (using structured prompts designed by the researchers) scored higher on clinical reasoning cases than physicians working through the same cases with access to the same LLM chatbot.^11,12^ It is unknown whether the difference in scoring could be attributed to different styles of inputting content into the LLM chatbot.

## OBJECTIVE

In this study, we aimed to identify how physicians interacted with an LLM chatbot (ChatGPT-4) in medical reasoning tasks to create a typology of input approaches, understand the rationales for different input approaches, and explore if any input practices were associated with improved clinical reasoning performance.

## METHODS

### Design

We carried out a sequential mixed methods study. A typology approach, which groups participants into types based on common features,^13^ was selected to explore different input styles. First, qualitative interviews were conducted with physicians who had completed mock clinical cases using ChatGPT-4 to understand input approaches and generate an initial typology. Next, the typology was used to analyze the recorded interactions (“chat logs”) between ChatGPT-4 and physician participants from the intervention arms of two randomized clinical trials (RCTs),^12,14^ where participants completed diagnostic and management cases using ChatGPT-4. Each clinical case where ChatGPT-4 had been used for problem-solving was coded to an input approach type. Lastly, we then examined whether the input approach types were associated with differences in performance by using a linear mixed-effects model to compare the scores and input approach types of the clinical cases. The design is visualized in Figure 1 and each research step is discussed below.

**Figure 1.**
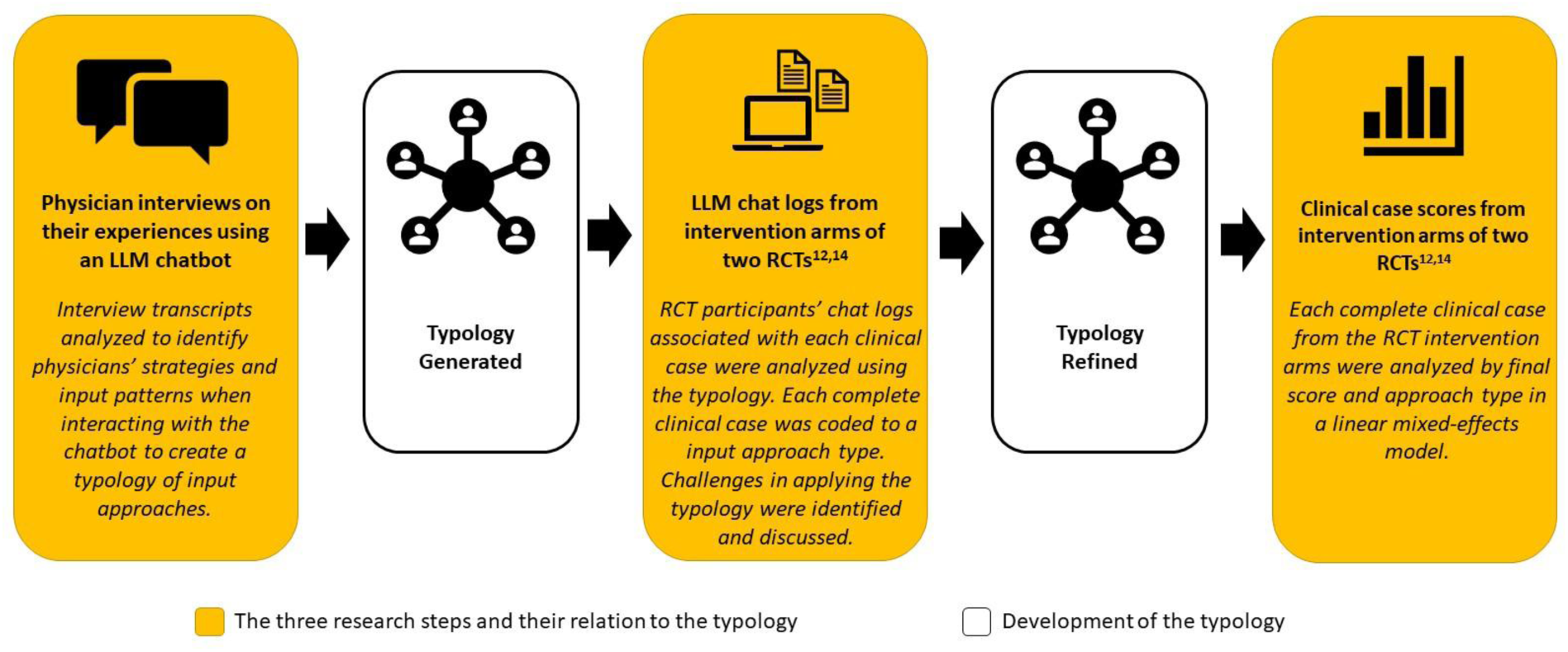
Steps in the development and application of the typology

This study underwent an exempt review by the Stanford Institutional Review Board, eProtocol #73932, and Beth Israel Deaconess Medical Center, Protocol #2024P000074. No AI tools were used for analysis or manuscript writing.

### Physician Interviews

We first conducted interviews with licensed physicians practicing in internal, family, or emergency medicine in the United States who were recruited by email from two previously conducted RCTs^12,14^ and by snowball sampling. Interested participants signed up for a 30-60 minute interview by either Google form or an email to the interviewer. Participants were offered an incentive of either $150 (for residents) or $199 (for attendings). Participants were first sent a Qualtrics survey comprised of three clinical case vignettes (two diagnostic and one management) to complete using ChatGPT-4 (see Appendix A). The diagnostic cases asked participants to present their top three differential diagnoses, data for and against each, and up to three suggested next steps in the diagnostic evaluation. The management case asked participants to describe the next appropriate step for management decisions, such as medication choices or conversations with patients and family. We then conducted qualitative semi-structured interviews with the participants. The interview topic guide (see Appendix B) covered the following topics: prior experience using AI tools, experiences using the chatbot to complete the mock cases, how they entered information into the chatbot, rationales for what they entered into the chatbot, and how they used the chatbot’s outputs in their decision-making. Interviews were conducted over Zoom by either LMH, an experienced qualitative health services researcher, or HK, a physician with training in conducting qualitative interviews. Interviews were recorded with permission and transcribed for analysis. Interviewers met regularly during data collection to discuss impressions, and continued conducting interviews until they agreed that no new themes arose.

Interview transcripts were imported into NVivo and analyzed using the Framework Method.^15,16^ The Framework Method is a structured approach to analysis using both deductive and inductive coding. All qualitative analysts (LMH, HK, RS) familiarized themselves with the interviews by listening to or reading the transcripts. Transcripts were coded by RS and initial coding was reviewed by LMH, with emergent codes reviewed by the entire qualitative team at monthly meetings and refined through discussion. After coding, a framework matrix was created in NVivo with participants represented in rows and code groupings presented as columns. Data was then charted into the framework matrix by summarizing the data within each cell to facilitate the identification of patterns of ChatGPT use, experiences using ChatGPT to work through the clinical cases, and participants’ rationales for different input strategies. For developing the typology, RS and LMH independently analyzed the described characteristics of inputs to develop categories of use, and then met to compare results and check for alignment before finalizing the typology.

### LLM Chat Log Coding

The recorded interactions between the intervention arm participants and the chatbot (chat logs) were downloaded from the two RCTs.^12,14^ In the intervention arms of each RCT, physicians could use ChatGPT-4 (access provided by the researchers) in addition to usual clinical care resources (e.g. UptoDate, Lexicomp) to complete clinical vignettes.^12,14^ Participants had a one-hour time limit and were instructed to prioritize quality of responses over completing all cases. In the diagnostic RCT, participants completed up to 6 cases,^12^ and in the management RCT, participants completed up to 5 cases.^14^

Three physician coders independently applied the definitions of input approach typologies to the chat logs, coding each completed clinical case to one of the input approach types in the framework, or as a new type if none applied. Cases, not participants, were coded to an approach type because physician participants may have used different approaches for different cases. After each chat log was coded by at least two physicians, the coding was reconciled. Any discrepancies in coding were discussed by the coders and the third physician acted as adjudicator as needed. Following chat log coding, the coders met to discuss coding challenges and check if there were any new approach types identified in the chat logs to further refine the typology.

### Clinical case scores and input approach type

In the two RCTs, each clinical case had undergone a grading process with multiple physician reviewers to determine a final score for that case.^12,14^ We then used a linear mixed-effects model to assess how each input approach type performed on diagnostic and management reasoning tasks by comparing the input approach and final scores for each case. Random effects for participant and case were included in the model to account for clustering by physician participant and case. A term for study, diagnostic, or management reasoning, was also included.

## RESULTS

We first present the findings from our interviews and the typology, followed by the analysis of input approach type performance.

### Participant characteristics

Twenty-two physicians were interviewed between May and July 2024; participant characteristics are described in Table 1. Interviews lasted a median of 33 minutes (range 16-54 minutes). Physicians reported different experiences using LLM chatbots prior to this study, with most having some experience using LLM chatbots, and six having no or minimal prior experience. Physicians may have used LLM chatbots in multiple ways (e.g., both personally and professionally), but were categorized in Table 1 according to the “highest” (most specialized) use case they described. Ten physicians described their LLM chatbot experience as self-taught, where their current knowledge came from reading information online or personal experimentation as opposed to formal training.

**Table 1:** Participant Characteristics.

| **Characteristic** | **N** |
| --- | --- |
| **Gender** |  |
| Female | 12 |
| Male | 10 |
| **Race/Ethnicity** |  |
| White | 12 |
| Asian | 3 |
| Asian: Indian | 3 |
| Hispanic | 3 |
| Native American | 1 |
| **Specialty** |  |
| Internal Medicine | 13 |
| Family Medicine | 5 |
| Emergency Medicine | 4 |
| **Years of Experience** |  |
| 1-3 | 15 |
| 4-9 | 5 |
| 10+ | 2 |
| **Type of Health System** |  |
| Academic Medical Center | 16 |
| County Hospital or Federally Qualified Health Center | 4 |
| Veterans Affairs | 1 |
| Private Practice | 1 |
| **Highest Level of Experience with Chatbot AI** |  |
| Clinical (e.g., differential diagnosis) | 5 |
| Professional (e.g., teaching, coding) | 8 |
| Personal (e.g., vacation planning) | 3 |
| None or minimal (e.g., “tried it once”) | 6 |

### Developing and refining the typology

From the interviews, we identified two distinct axes of inputting information into the chatbot. The first axis is “content amount.” Physicians described inputting four different amounts of content from the vignette, from “most” to “least”: copy-pasting the entire vignette, copy-pasting sections of the vignette, summarizing the vignette in their own words, or writing brief queries about topics in the vignette. The second axis is “prompt style” which involved telling the chatbot what to do with the information, such as providing context (e.g., “you are helping a physician solve a clinical case”) or asking it to perform a task (e.g., “provide a differential diagnosis”). In the analysis of chat logs from the RCTs, coders found that the descriptions for “content amount” could be reliably identified, with a predominant input approach used for most clinical cases. However, “prompt style” was difficult to identify since physicians may have tried different types of prompts within a single case. Because prompt style was not clearly identifiable within the chat logs, the final typology was refined to address content amount only, and is described in Table 2.

**Table 2:** Typology of Input Approaches.

| Type | Description | Exemplary Quote |
| --- | --- | --- |
| Copy-paster | Copies and pastes the whole vignette into the chatbot. | *I basically just inputted the entire vignette and then said, "Please list three potential diagnoses based off of the vignette." Physician 14*<br>*If I typed every single thing that I thought was important, that was going to take a lot more time than just me copying everything and just pasting it. Physician 22*<br>*It's definitely less effort to just copy-paste the whole thing in [...] instead of carefully formulating a well-thought-out question. Physician 10*<br>*I think [copy-pasting] was just to give it all the information that we were provided with. I wasn't sure if there's a better way to format it. Physician 13* |
| Selective copy-paster | Copies sections of the vignette, but not the whole thing; makes selections of content to copy based on what is believed to be relevant. | *I mostly did chunks of [the vignette]. I usually just did the descriptive parts. Physician 3*<br>*I might copy and paste the relevant part of the physical exam, but not the whole physical exam [...] I try to narrow the search space so that I'm really getting what I want back. Physician 16*<br>*I just had this assumption it would just overwhelm ChatGPT. So I didn't copy and paste the whole case. I just copy and pasted more of the aspects of the information that would be more valuable to me in terms of affecting my decision making. Physician 15* |
| Summarizer | Makes assessment of relevant or important information from the vignette and summarizes the information in their own words to enter into the chatbot. | *I only put the relevant information, or at least what looked like the relevant information to me, from a case. I would summarize everything in there. Physician 16*<br>*I more often tend to just summarize the pertinent findings. [...] I feel like just convenience for myself, and also trying to reduce the amount of noise for ChatGPT. I don't want it going down rabbit holes. Physician 19*<br>*Another time I tried writing the case with the points that I thought were pertinent positives. I tried to distill it to, "All right, these are the pertinent positives." [...] I don't want to feel like I'm outsourcing all of my thinking and literally just copy-pasting. Physician 17* |
| Searcher | Uses the chatbot like internet search with basic queries; might include stringing together multiple concepts to ask a question, but limited to no more than one sentence. | *There were specific questions that I typed in the ChatGPT, because I needed the answer to come up with the three differential diagnoses. Physician 8*<br>*I wanted to use it more as a tool to supplement me if I needed it. And that's why I only asked it, 'I'm thinking this, this is what I'm going to put in.' [...] I wanted to see if the data supported that and not have a computer give me a diagnosis initially. Physician 2*<br>*I tried a couple more generic questions and the responses I got back were I think too vague to be helpful. [...] I have a hard time knowing how to communicate what I need out of ChatGPT to give me back something useful. Physician 6* |

### Physicians’ experiences using the chatbot by type

Physicians were asked to describe their rationale for inputting content into the chatbot and their perceptions of the subsequent outputs. These are explored in the next sections, organized by the approach taken for each case. Of note, nine interviewed physicians described trying more than one approach, as opposed to consistently using one strategy across all three cases. They described trying different approaches either because they wanted to experiment with seeing what the chatbot would do with a different amount of content, or because they felt that some cases would be better approached with a different input strategy. Some physicians who had not used LLM chatbots prior to the interviews described “discovering” that they could copy and paste large amounts of content.

#### Copy-Pasters

Fifteen interviewed physicians described copy-pasting the entire vignette for at least one case. One rationale given for utilizing copy-pasting was in how it was perceived as the “easiest” method, either in how copy-pasting took less time than typing out new text, or for how using the instructions embedded in the case (i.e., “list three possible diagnoses”) was simpler than reformatting a new query. Some physicians also described utilizing copy-pasting when they wanted a more robust response from the chatbot, based on perceptions that a more detailed input would produce a more detailed output. Copy-pasting for the diagnostic cases was generally found to produce a detailed differential that could be quickly skimmed, and was described as particularly useful for idea generation or confirmation. One physician described this as a “shotgun” approach, where they felt that a comprehensive list of possible diagnoses was an efficient way to “get their brain working.” In contrast, two physicians thought the outputs were too verbose to be very helpful to their problem-solving process.

#### Selective Copy-Pasters

Six physicians described using selective copy-pasting for at least one case. Selective copy-pasting allowed physicians to quickly capture specific pieces of the vignette that were of interest. Physicians described two primary rationales: 1) to focus only on specific pieces of the vignette, such as the labs or symptom description, or 2) because they were unsure of how much content the chatbot could “handle” and assumed less content would avoid “overwhelming” it. The outputs were generally described as helpful, but three physicians who experimented with copy-pasting on subsequent cases noted that by comparison, selective copy-pasting was a slower process and seemed to produce less detailed outputs.

#### Summarizers

Four physicians described using summarizing for at least one clinical case. Similar to selective copy-pasting, the rationale was to create an input that captured details of interest while leaving out information perceived as irrelevant. In contrast to selective copy-pasting, writing out a summary was perceived as a more flexible approach as physicians could incorporate information across the vignette in their own words, focusing only on the information they felt was most relevant to prevent the chatbot from “going down rabbit holes.” In contrast, one physician tried summarizing primarily because they disliked how copy-pasting felt too much like “outsourcing” all of their thinking to a chatbot. For this participant, the process of creating their own summary helped them feel more cognitively engaged in the problem-solving process. One limitation of summarizing was that information could sometimes be missed, which was noted by a physician who had also tried copy-pasting and felt that the chatbot picked up on aspects of the case that they had “glossed over” in their summary.

#### Searchers

Seven physicians described using searching by inputting brief queries, similar to how one would use a search engine, for at least one case. Not every physician had a specific rationale for utilizing searching—one physician was unsure, and presumed that it may be because it was how they were used to looking up information on the internet. Some physicians noticed that large inputs would create large outputs, and felt that a more specific query was the best method for getting a concise output that addressed only their question and did not contain “irrelevant” information that they would have to take time to read through. Searching was often used when the physician already had a clear idea of how they were going to answer the case, but only needed only a few pieces of specific information to complete their answer. The same physician who described copy-pasting as a “shotgun” approach also tried searching, and described this as a “sniper approach” that was valuable when seeking an answer for a “targeted” or specific question. Most physicians felt that they were easily able to elicit useful outputs based on their inputs. However, two physicians experienced more difficulty in generating relevant outputs, finding it challenging to design the right question for the chatbot.

### Input approach type and clinical decision-making performance

Figure 2 depicts the scores of each participant case from the intervention (access to the chatbot) arms of both RCTs, plotted by their type. In the diagnostic RCT, 19 physicians completed a total of 95 diagnostic cases that had both an accessible chatlog and a score. In the management RCT, 42 physicians completed a total of 158 cases with both an accessible chatlog and a score. In the original RCTs, 10 additional physicians participated (6 in the diagnostic RCT; 4 in the management RCT), but the chat logs were lost and could not be downloaded, and so were excluded from this analysis. Cases that were started but not completed (e.g., had a chat log but were given a score of 0) were not included in the analysis.

**Figure 2.**
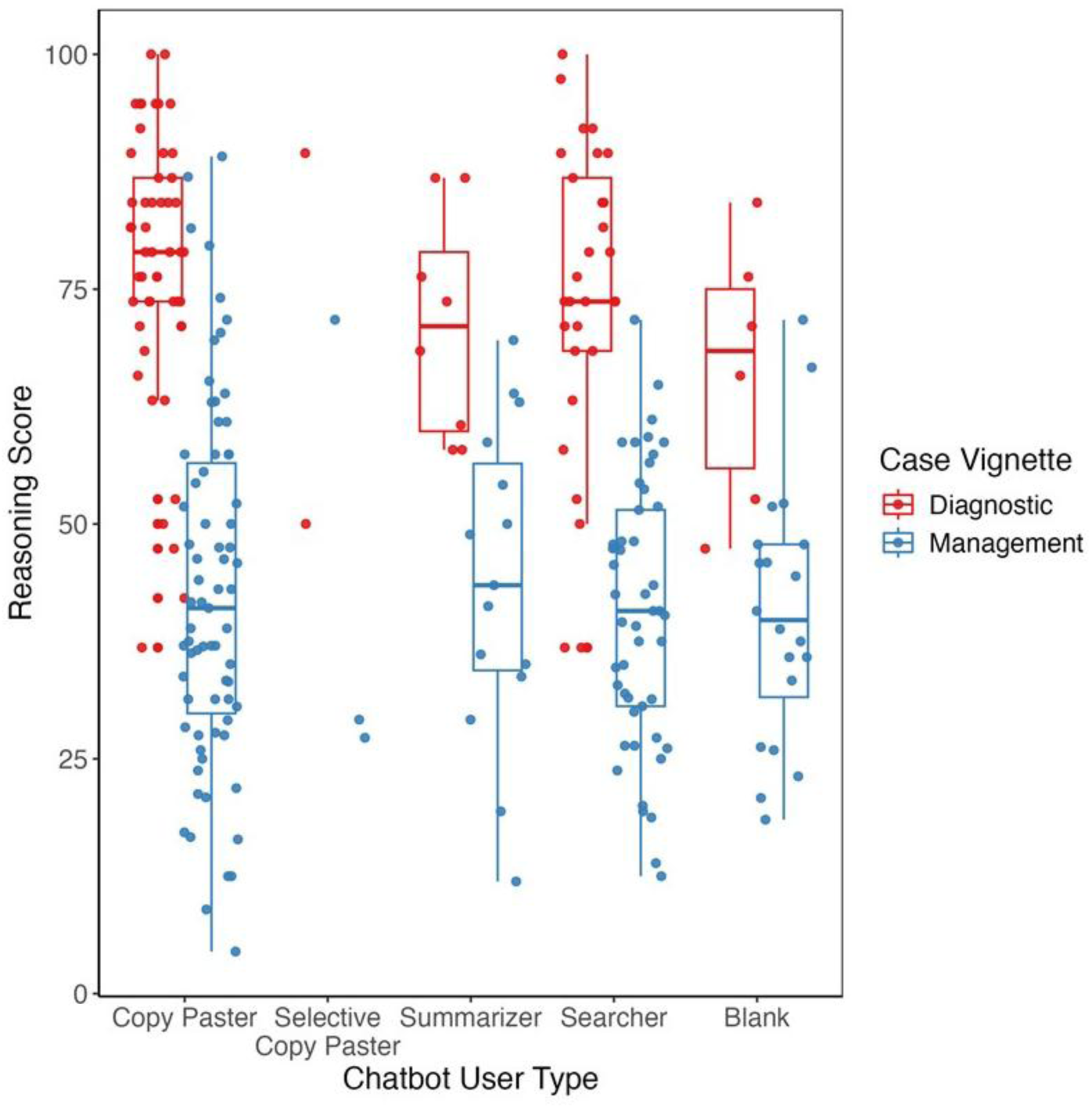
Clinical reasoning scores for diagnostic and management cases organized by content type. Each point represents a participant’s score on a vignette case. Analyzed using a linear mixed-effects model with random effects for participant and case to account for clustering by physician participant and case

All four types were represented in both the diagnostic and management RCTs, but most participants worked through the cases by either copy-pasting or searching. In some instances, seen more often with the management cases, a case was worked through and completed without using the chatbot at all (plotted in the “blank” column). No single type was associated with scoring higher on the clinical cases, for both diagnostic and management cases.

## DISCUSSION

In this study, we developed a typology of how physicians interacted with an LLM chatbot and explored if input practices were associated with improved clinical reasoning. As LLM chatbots grow in popularity and demonstrate greater abilities in working through clinical cases, this study adds to our understanding of how physicians approach using LLM chatbots and what may be needed to effectively collaborate with chatbots in clinical settings.

Our typology identified that physicians utilized four distinct “content amount” input styles (copy-pasting, selective copy-pasting, summarizing, and searching) when entering information into the chatbot. While no single style was associated with higher scores on the clinical cases, copy-pasting and searching were the approaches utilized most. Previous studies of lay people’s use of chatbots for working through word problems have also reported frequent use of either copy-pasting or searching, where copy-pasting was often selected when users perceived copying as easier, while searching was often selected due to prevailing mental models of search engines.^17,18^ Some physicians who used searching also frequently described aiming for more narrow outputs, whereas some physicians who used copy-pasting described aiming for more robust outputs. The literature on clinical problem-solving has highlighted how experienced physicians can vary greatly on the amount or type of data needed to work through the same clinical cases, which may be due to personal problem-solving styles or the uniqueness of their knowledge bases.^19,20^ This typology elucidates the different intuitions and intentions that physicians may have when interacting with an LLM chatbot in the context of clinical problem-solving, which may be useful for either developing training on LLM chatbot usage that is tailored to physicians, or for developers to design LLM chatbots that are aligned with how physicians approach using digital tools for clinical problem-solving.

Previous research has highlighted the significant potential that LLM chatbots may have in supporting clinical decision-making, such as through retrieving medical knowledge and generating differential diagnoses and treatment plans.^21^ However, our results also highlight how the user will need to be able to generate relevant information efficiently in order for these tools to be useful within the decision-making process. Some of the physicians interviewed in this study described getting frustrated when they were unable to get the desired response from the chatbot, and a few felt that they may have been faster working on their own. And in the coding of the RCT chat logs, some were “blank,” meaning that physicians did not use the chatbot at all while working through a case, which occurred more often for management cases. In the interviews, physicians universally felt that the outputs for management cases were less helpful than the diagnostic cases, describing them as too broad and unable to consider patient- and setting-specific nuances. Many stated that they relied more on their own clinical experience to work through these cases, which may be one reason why the chatbot was not utilized as much for the management cases. Barmen et al. (2024) have argued that to support LLM users, there should be guidelines clarifying which tasks LLM chatbots can and can’t do well, which tasks require additional refinement, and “context-specific heuristics.”^22^ It will be important to continue to disseminate trainings and information on the different available LLM chatbot tools and their abilities, the strengths and limitations in using LLM chatbots for different clinical tasks, and how to best tailor LLM chatbot use to a case, patient, or setting to support physicians in using these tools.^26^

Our study was novel for seeking to understand how physicians working through clinical cases would intuitively approach selecting inputs for chatbots. Though the “prompt style” axis was not included in the final typology, asking the chatbot to “do something” with the content provided was still highly utilized by physicians, who used a range of different prompting styles both across and within cases, with some even describing having a “conversation” with the chatbot and asking it to provide reasoning. The results of our analysis of clinical case score by content amount type showed both a range of scores within each type, and also that no single type performed better than others. It may be the case that content amount alone does not significantly impact the accuracy or utility of outputs. Research continues to test the effectiveness of different prompting techniques in the context of clinical problem-solving.^6,23^ Guides for medical professionals have identified various best practices, such as assigning a role, providing context, requesting examples, giving instructions, or specifying the desired output.^24,25^ It may be the case that structured prompt engineering techniques such as these have more impact on the quality of the final output than the amount of content provided alone. It will be important for future research to continue to examine prompt engineering strategies and effectiveness in generating accurate, useful outputs when using chatbots for clinical cases, as well as how best to educate physicians in their use. Since our results show that organic input styles by physicians can be quite variable, we suggest that future research first test prompting styles in controlled settings to identify which strategies are most effective before testing their effectiveness with real-world users.

## LIMITATIONS

All cases were based on real patient encounters, but did not involve real patients and were presented as vignettes. Participants from the two RCTs also had a time limit for completing the clinical cases. Participants may have approached information gathering and problem-solving in a real-world scenario differently, yet these mock cases are still informative, helping us understand how physicians might approach using a LLM chatbot for problem-solving. This study also involved a large number of resident trainees, but only licensed second-year residents and above were recruited to ensure participants had sufficient experience. This study mostly involved inpatient practitioners, and the experiences of outpatient practitioners should be explored in future studies. There is high risk of confounding since strategy choice was not random and participants likely chose specific strategies based on factors related to case difficulty and their own knowledge in the clinical content area. We did not examine the chat log outputs for accuracy or quantity of content and so could not assess the variability in the chatbot responses, though LLMs are known to provide different responses to the same question.

## CONCLUSION

Physician usage and understanding of LLM chatbot AI systems is variable, from copy-pasting entire clinical cases to treating chatbot systems similarly to search engines with brief queries. Purposeful training and support is needed to acclimate physicians to learn how to effectively use emerging AI technologies to realize their potential for supporting safe and effective medical decision-making in practice.

## Data Availability

All data produced in the present study are available upon reasonable request to the authors.

## ETHICS APPROVAL

This study was conducted in accordance with federal and state regulations and ethical guidelines for biomedical research, and underwent an exempt review by the Stanford Institutional Review Board, eProtocol #73932, and Beth Israel Deaconess Medical Center, Protocol #2024P000074.

## COMPETING INTERESTS

Jason Hom reports

- Being an advisor for Cognita Imaging and having equity options

Jonathan Chen has received additional research funding support in part by

- NIH/National Institute of Allergy and Infectious Diseases (1R01AI17812101)
- NIH-NCATS-Clinical & Translational Science Award (UM1TR004921)
- NIH/National Institute on Drug Abuse Clinical Trials Network (UG1DA015815 - CTN-0136)
- Stanford Bio-X Interdisciplinary Initiatives Seed Grants Program (IIP) [R12] [JHC]
- NIH/Center for Undiagnosed Diseases at Stanford (U01 NS134358)
- Stanford Institute for Human-Centered Artificial Intelligence (HAI)

Jonathan Chen reports being

- Co-founder of Reaction Explorer LLC that develops and licenses organic chemistry education software.
- Paid medical expert witness fees from Sutton Pierce, Younker Hyde MacFarlane, Sykes McAllister, and Elite Experts.
- Paid consulting fees from ISHI Health.
- Paid honoraria or travel expenses for invited presentations by General Reinsurance Corporation, Cozeva, and other industry conferences, academic institutions, and health systems.

Dr Rodman reports additional research funding paid in part by:

- Josiah Jr. Macy Foundation (P25-04)

Dr. Rodman reports being

- A part-time visiting researcher at Google

## REFERENCES

1. Hu K. ChatGPT sets record for fastest-growing user base - analyst note [Internet]. Reuters. 2023 [cited 2024 Dec 16];Available from: https://www.reuters.com/technology/chatgpt-sets-record-fastest-growing-user-base-analyst-note-2023-02-01/

2. Kisvarday S, Yan A, Yarahuan J, et al. ChatGPT Use Among Pediatric Healthcare Providers (Preprint). JMIR Form Res 2024;

3. Eppler M, Ganjavi C, Ramacciotti LS, et al. Awareness and Use of ChatGPT and Large Language Models: A Prospective Cross-sectional Global Survey in Urology. Eur Urol 2024;85(2):146–53.

4. Strong E, DiGiammarino A, Weng Y, et al. Chatbot vs Medical Student Performance on Free-Response Clinical Reasoning Examinations. JAMA Intern Med 2023;183(9):1028–30.

5. Lahat A, Sharif K, Zoabi N, et al. Assessing Generative Pretrained Transformers (GPT) in Clinical Decision-Making: Comparative Analysis of GPT-3.5 and GPT-4. J Med Internet Res 2024;26(1).

6. Wang L, Chen X, Deng XW, et al. Prompt engineering in consistency and reliability with the evidence-based guideline for LLMs. NPJ Digit Med 2024;7(1).

7. Zhang C, Zhang C, Li C, et al. One Small Step for Generative AI, One Giant Leap for AGI: A Complete Survey on ChatGPT in AIGC Era. 2023;Available from: http://arxiv.org/abs/2304.06488

8. Muktadir GMD. A Brief History of Prompt: Leveraging Language Models. (Through Advanced Prompting). arXiv:231004438 [Internet] 2023;Available from: https://www.researchgate.net/publication/372830312

9. Chen C, Lee S, Jang E, Sundar SS. Is Your Prompt Detailed Enough?: Exploring the Effects of Prompt Coaching on Users’ Perceptions, Engagement, and Trust in Text-to-Image Generative AI Tools. In: ACM International Conference Proceeding Series. Association for Computing Machinery; 2024.

10. Zamfirescu-Pereira JD, Wong RY, Hartmann B, Yang Q. Why Johnny Can’t Prompt: How Non-AI Experts Try (and Fail) to Design LLM Prompts. In: Conference on Human Factors in Computing Systems - Proceedings. Association for Computing Machinery; 2023.

11. McDuff D, Schaekermann M, Tu T, et al. Towards Accurate Differential Diagnosis with Large Language Models. 2023;Available from: http://arxiv.org/abs/2312.00164

12. Goh E, Gallo R, Hom J, et al. Large Language Model Influence on Diagnostic Reasoning. JAMA Netw Open [Internet] 2024;7(10):e2440969. Available from: https://jamanetwork.com/journals/jamanetworkopen/fullarticle/2825395

13. Stapley E, O’Keeffe S, Midgley N. Developing Typologies in Qualitative Research: The Use of Ideal-type Analysis. Int J Qual Methods 2022;21.

14. Goh E, Gallo RJ, Strong E, et al. GPT-4 assistance for improvement of physician performance on patient care tasks: a randomized controlled trial. Nat Med 2025;

15. Gale NK, Heath G, Cameron E, Rashid S, Redwood S. Using the framework method for the analysis of qualitative data in multi-disciplinary health research. BMC Med Res Methodol 2013;13(1):117.

16. Ritchie J, Lewis J, editors. Qualitative Research Practice: A Guide for Social Science Students and Researchers. London: SAGE Publications; 2003.

17. Krupp L, Steinert S, Kiefer-Emmanouilidis M, et al. Unreflected Acceptance -- Investigating the Negative Consequences of ChatGPT-Assisted Problem Solving in Physics Education. 2023;Available from: http://arxiv.org/abs/2309.03087

18. Khurana A, Subramonyam H, Chilana PK. Why and When LLM-Based Assistants Can Go Wrong: Investigating the Effectiveness of Prompt-Based Interactions for Software Help-Seeking. In: ACM International Conference Proceeding Series. Association for Computing Machinery; 2024. p. 288–303.

19. Jones RH. Data collection in decision-making: a study in general practice. Med Educ 1987;21(2):99–104.

20. Nendaz MR, Gut AM, Perrier A, et al. Common strategies in clinical data collection displayed by experienced clinician-teachers in internal medicine. Med Teach 2005;27(5):415–21.

21. Vrdoljak J, Boban Z, Vilović M, Kumrić M, Božić J. A Review of Large Language Models in Medical Education, Clinical Decision Support, and Healthcare Administration. Healthcare (Switzerland). 2025;13(6).

22. Barman KG, Wood N, Pawlowski P. Beyond transparency and explainability: on the need for adequate and contextualized user guidelines for LLM use. Ethics Inf Technol 2024;26(3).

23. Patel D, Raut G, Zimlichman E, et al. Evaluating prompt engineering on GPT-3.5’s performance in USMLE-style medical calculations and clinical scenarios generated by GPT-4. Sci Rep 2024;14(1).

24. Meskó B. Prompt Engineering as an Important Emerging Skill for Medical Professionals: Tutorial. J Med Internet Res 2023;25(1).

25. Shah K, Xu AY, Sharma Y, et al. Large Language Model Prompting Techniques for Advancement in Clinical Medicine. J Clin Med. 2024;13(17).

