## Appendix A for "A typology of physician input approaches to using AI chatbots for clinical decision-making: a mixed methods study"

### Intro Block

Thank you for participating in our research study. Throughout the survey, you will work on **2 diagnostic cases and 1 management case**, presented in a randomized order.

Before we proceed, please kindly provide your information.

First and Last Initials followed by home street number
Ex: if your name was Tom Ford and you live at 1111 Main Street, you would put “TF1111”).

Email

Specialty

- Internal Medicine

- Family Medicine
- Emergency Medicine

How many years of experience in medicine do you have, including residency training?
Ex: if you have completed 3 years of residency training and have been an attending for two years, then you would enter "5".

Prior Experience with Generative AI / ChatGPT

- I've never used it before
- I've used it once ever
- I use it rarely (less than once per month)
- I use it occasionally (more than once per month but less than weekly)
- I use it frequently (weekly or more)

**Diagnostic Case Example**

### Example:

In this study, you are going to read diagnostic cases and complete three discrete sections for each case: structured reasoning, your final diagnosis, and the next diagnostic steps. Here is an example case.

### Example Case

**HISTORY OF PRESENT ILLNESS**
A 24-year-old woman who is sixteen weeks pregnant is seen in the office twelve hours after the onset of abdominal pain. The pain is severe, steady, and located in the epigastrium. She vomits several times during the interview. She does not have diarrhea. Her pregnancy has been uneventful. She has no history of gastrointestinal disease. Her only medications are a daily multivitamin

**PHYSICAL EXAMINATION**
On examination the blood pressure is 110/60 mmHg and the pulse 88/min. and regular. The temperature is 38.2°C. Mild jaundice is present, but there are no spider angiomas. The eyes and nose are normal, but the tympanic membranes show no light reflex. The cardiac PMI is normally placed, and there is a soft systolic murmur along the left sternal border. The second heart sound splits normally. Severe epigastric tenderness is present and the bowel sounds are hypoactive. The liver is not palpable, but the spleen is two fingerbreadths below the left costal margin. The uterus is palpable 10 cm above the pubis.

**LABORATORY EXAMINATION**
Hematocrit 39%
WBC count 12,600/mm3
Platelets 185,000/mm3
Neutrophils 88%
Bands 3%
Lymphocytes 9%
Bilirubin (total) 4.6 g/dL
Alkaline phosphatase 300 U/L

ALT 120 U/L
AST 100 U/L
Albumin 3.2 g/dL
Total protein 6.5 g/dL
Prothrombin time, normal
Amylase 860 U/L
Lipase 1210 U/L
pH 5.6 (urinalysis)
RBCs 0/HPF (urinalysis)
WBCs 2-3/HPF (urinalysis)
Bacteria occasional/HPF (urinalysis)

**RADIOLOGY**
A plain film of the abdomen shows a normal bowel gas pattern.

### Example Matrix (Answer)

**PART 1**: Structured Reasoning

| | Diagnosis<br><br>List 3 possible diagnoses below | Supporting Factors<br><br>For each possible diagnosis listed, provide findings/risk factors supporting this hypothesis | Opposing Factors<br><br>For each possible diagnosis listed, provide findings opposing this hypothesis, or findings that were expected by not present |
|---|---|---|---|
| 1 | Gallstone pancreatitis | Elevated Lipase, Amylase, Bilirubin, AST, ALT, WBC, Temp, neutrophils, bands. Vomiting + epigastric pain. | No pain with eating. Young age for gallstones |
| 2 | Pregnancy Loss / Spontaneous Abortion | Pain (cramps), known pregnancy | No vaginal bleeding. Many unexplained lab abnormalities including elevated lipase, AST/ALT |
| 3 | HELLP Syndrome | Pregnancy, transaminitis<br>Abdominal pain | Early in pregnancy<br>No hypertension or proteinuria to suggest preeclampsia. No hemolysis or low platelets |

Example Most likely Diagnosis

**PART 2: Final diagnostic decision**

Based upon your reasoning above, what is your final diagnosis?

Gallstone Pancreatitis

Click to write the question text

**PART 3: Additional Steps**

Name up to 3 additional steps that you would take in your diagnostic process

| | |
|---|---|
| 1 | Ultrasound of the abdomen including right upper quadrant and uterus |
| 2 | Additional laboratory tests: fractionated bilirubin, Calcium, triglycerides |
| 3 | Ask about amount and pattern of alcohol intake as a risk factor for alcoholic pancreatitis. |

### Diagnostic Case 1

### DIAGNOSTIC CASE 1 (3 Parts)

#### History Of Present Illness

A 76M comes to his PCP complaining of pain in his back and thighs for 2 weeks. He has no pain sitting or lying, but walking causes severe pain in his low back, buttocks and calves. He feels febrile and tired. He was told by the referring cardiologist that his recent tests results since the pain started showed a new anemic and azotemia. A few days before the onset of the pain he had undergone coronary angioplasty. Heparin was administered for 48 hours.

#### Past Medical History

Ischemic heart disease had first been diagnosed ten years earlier, at which time a coronary artery bypass procedure was done.

**Physical Examination**

VITALS: 99.6° F.; pulse was 94/min and regular; BP was 110/88 mmHg.
GEN: Well appearing
CARDS: There is a grade III/VI apical systolic murmur; lungs were clear to auscultation.
PULM: Lungs are clear to auscultation bilaterally, no wheezing, or consolidations noted
ABD: Soft, non-tender to palpation
MSK: He does not have tenderness of his spine or pelvis. Spinal mobility is normal, as is the mobility of his hips. Standing is painless; however, pain is experienced in his low back, buttocks and calves within a minute of feeble running in place. The pain disappears shortly after exercise is discontinued.
EXT: Peripheral pulses were symmetrically reduced, but palpable. The neurological examination was normal.
SKIN: Patient has a purple, red, lacy rash over his low back and buttocks.

**Laboratory**

WBC of 11.5 x 103 cells /μL; differential of 64% segs, 20% lymphocytes, 3% monocytes, 12% eosinophils and 1% basophil. The hematocrit was 28% and the platelet count was 315 x 103 /μL. The erythrocyte sedimentation rate was 99 mm/h. Urinalysis was normal except for 2+ proteinuria. Serum creatinine was 4.0 mg/dL; sodium was 145 mEq/L; potassium 4.0 mEq/L; chloride 105 mEq/L. SGOT was 27 U/L; GGT was 90 U/L; alkaline phosphatase was 153 U/L.

**PART 1**: Structured Reasoning

| | Diagnosis<br><br>List 3 possible diagnoses below | Support diagnosis<br><br>For each possible diagnosis listed, provide findings/risk factors supporting this hypothesis | Opposing diagnosis<br><br>For each possible diagnosis listed, provide findings opposing this hypothesis, or findings that were expected by not present |
|---|---|---|---|
| 1 | | | |
| 2 | | | |
| 3 | | | |

**PART 2: Final diagnostic decision**

Based upon your reasoning above, what is your final diagnosis?

**PART 3: Additional Steps**

Name up to 3 additional steps that you would take in your diagnostic process

1

2

3

### Diagnosis Case 2

### DIAGNOSTIC CASE 2 (3 Parts)

#### History of Present Illness

A previously healthy 64M presents with a six month history of fatigue, 30 pound weight loss, and worsening skin rash. He states, "I have no energy." In the past month, he suddenly cannot climb stairs or rise from a chair without assistance. The rash, which began on his elbows and knees, has spread to his hands and face. He has minimal appetite, but feels constantly bloated and notes constipation He denies pain, hoarseness, or dysphagia. His only medication is topical steroids recently provided for his skin rash.

#### Past History

Until this illness he developed the skin rash he had never seen a physician.

#### Systems Review

His system review is negative except for the present illness.

#### Family History

Father died at 64 with colon cancer. Mother died at 60 from a stroke. One brother died at 66 with renal cell carcinoma.

**Social History**
He smokes one pack of cigarettes per day. He drinks two "six-packs" most weekends.

**Physical Examination**
VITALS: His blood pressure is 140/90 mmHg, his pulse is 80/min., and his respiratory rate is 15/min.
GEN: The patient is a thin, pale, white male in no acute distress.
CARD: Normal rate and rhythm and no murmurs, gallops, rubs
PULM: Lungs are clear to auscultation bilaterally
ABD: The abdomen is distended and is tympanitic to percussion. Bowel sounds are normal. The rectal exam reveals no masses, but the stool is positive for occult blood
GU: The GU exam is normal.
EXT: No pitting edema
Skin: scaly, erythematous eruption involving the dorsum of the elbows, knees, and interphalangeal and metacarpophalangeal joints. A blue-red rash is present on his face
Neuro: Neurological examination revealed symmetrical proximal quadriceps muscle weakness of the iliopsoas and quadriceps

**Laboratory and Other Procedures**
WBC is 12,400/μL with 82% PMN's, 16% lymphocytes and 2% monocytes. Hemoglobin is 9.5 g/dL, hematocrit 30% and MCV is 69 fL. Platelets are 660,000/μL. The erythrocyte sedimentation rate is 90 mm/hr. Chemistry panel: Normal except for the Creatine phosphokinase which is 440 IU (nl 47-220).

**PART 1**: Structured Reasoning

| | Diagnosis<br><br>List 3 possible diagnoses below | Support diagnosis<br><br>For each possible diagnosis listed, provide findings/risk factors supporting this hypothesis | Opposing diagnosis<br><br>For each possible diagnosis listed, provide findings opposing this hypothesis, or findings that were expected by not present |
|---|---|---|---|
| 1 | | | |
| 2 | | | |
| 3 | | | |

**PART 2: Final diagnostic decision**
Based upon your reasoning above, what is your final diagnosis?

**PART 3: Additional Steps**
Name up to 3 additional steps that you would take in your diagnostic process

1

2

3

### Management Case 1

**MANAGEMENT CASE (5 Questions)**
**HISTORY OF PRESENT ILLNESS:**
A 72 year-old Vietnamese-speaking woman with HLD, HTN, CKD stage II (baseline creatinine 1.3), cholelithiasis is admitted to the hospital for severe gallstone pancreatitis.

PMH/PSH:
Hyperlipidemia
Hypertension
Cholelithiasis
CKD stage II
History of positive PPD
Right knee replacement age 65

FHx:
Heart disease
Diabetes

SHx:
Former smoker
Drinks 1-2 drinks of ETOH daily

Lives with her son and daughter in law
Widowed
Used to work managing in a textile factory

Pre-admission medication list:
-Atorvastatin 40mg qhs
-Lisinopril 5mg qD
-Ibuprofen 400-800mg qD PRN for joint pain

**PHYSICAL EXAM**:
T 101.3, HR 112, BP 98/52, O2 sat 93% RA, RR 24
Gen: Alert, oriented, in NAD, appears very fatigued
HEENT: mild scleral icterus
CV: tachycardic, regular rhythm, no m/r/g
Lungs: crackles in the bases bilaterally
Abdomen: soft, mildly distended, TTP in the RUQ/peri-umbilical areas
Ext: no LE edema
MSK: no joint swelling

**LABORATORY AND OTHER PROCEDURES**:
CBC showed leukocytosis to 17.3K, decreased Hgb to 10.2, and elevated platelet count to 345K.
BMP showed elevated creatinine to 1.8, elevated BUN to 45.
LFTs show elevated AP to 102, elevated Tbili to 3.2.
Lipase is elevated to 603.
CT A/P shows peripancreatic stranding consistent with pancreatitis, cholelithiasis, and intrahepatic biliary ductal dilatation.

**CASE CONTINUATION**:
Her course is complicated by severe sepsis and ARDS requiring 6L supplemental O2, for which she is transferred to the ICU. She is treated with broad spectrum antibiotics and frequent electrolyte repletion. A chest x-ray obtained to evaluate for pneumonia shows a lung nodule, bilateral

interstitial markings but no consolidation. After transfer out of the ICU, she has a follow up CT with contrast that showed improvement of the interstitial edema and reconfirms a single 2.5 cm nodule; the radiology report suggests either biopsy or repeat imaging in three months. Because she immigrated from Vietnam and has a history of positive PPD, the infectious disease team is consulted and recommends ruling her out for tuberculosis. Three acid-fast bacilli smears are sent, as is a nucleic acid amplification test, and all are negative for tuberculosis.

Her course is further complicated by the development of a DVT, which requires a heparin drip with transition to apixaban, as well as respiratory failure requiring intubation. Despite a prolonged hospital course – she is an inpatient for 57 days – she gradually recovers. She is severely deconditioned, and rehabilitation is recommended.

On the day of discharge – 54 days after her lung nodule is discovered – she is preparing for discharge. Her lung nodule has long ago dropped off her progress notes, but it is again noted on a review of imaging prior to discharge.

**DISCHARGE MEDICATION LIST**:
-Atorvastatin 40 mg qhs
-Lisinopril 5 mg qD on HOLD until patient follows up with PCP
-Ibuprofen 400-800 mg qD PRN for joint pain on HOLD given mild AKI on CKD during hospitalization
-Acetaminophen 1000 mg q8h PRN for pain
-Potassium chloride 20mEq daily
-Apixaban 5mg BID

**DISCHARGE PHYSICAL EXAM**:
T 97.3F, HR 85, BP 142/75, O2 sat 95% RA, RR 18
Gen: Alert, oriented, in NAD, appears weak
HEENT: no scleral icterus, no oral lesions

CV: RRR, no m/r/g
Lungs: CTAB, no c/r/w
Abdomen: soft, non-distended, non-tender to palpation
Ext: no LE edema
MSK: no joint swelling

**DISCHARGE LABS**:
CBC:
WBC 10.5
Hgb 8.3
Plt 473

BMP:
Sodium 137
K 3.4
Chloride 104
Bicarb 21
BUN 26
Creatinine 1.5

**Question 1**

Based on the information you have at this time, what is the differential diagnosis for this incidental lung nodule?

**Question 2**

What additional information (from patient or medical record) could you non-invasively obtain that would help further hone the differential diagnosis for this lung nodule?

**Question 3**

This patient is being planned for discharge today and is expecting to leave today to go to rehab. What factors would influence your decision on keeping the patient in the hospital or deferring to the outpatient setting for additional work up?

**Question 4**

If you were to defer the workup to the outpatient setting, what would be your process?

### Question 5

If you were to keep the patient in the hospital for additional investigations, at what point in those investigations would you discharge the patient?

Powered by Qualtrics
