## Appendix B for "A typology of physician input approaches to using AI chatbots for clinical decision-making: a mixed methods study"

**Topic Guide**

**Introduction:**

This is a multi-center qualitative study assessing physicians' views on the use of artificial intelligence in clinical practice. We will be performing a series of interviews with around 20 doctors. All information we gather will be anonymized and securely stored.

Do you have any questions for me before we begin?

Is it OK to record?

| Main questions | Prompts |
|---|---|
| **Background**: First, I'd like to start with some background information. | |
| Could you tell me a bit about yourself, like your clinical interests and how long you've been in practice? | |
| What do you enjoy most about being a doctor? | |
| Do you have any previous experience using AI for either clinical or personal use? [ChatGPT, Dali, Claude, BingAI] | If yes, probe for brief details.<br>Have you had any training in creating prompts for working with AI? |
| **Views on GPT**: Thanks for that background information. I now want to talk about your experience using GPT for clinical decision making, as you did in the survey with clinical questions for this study. | |
| How did it go using GPT for clinical decision making? | How did you input information or questions to GPT? Why did you input information in that way?/ Why did you take the approach you did to using GPT?<br>What did you like about GPT?<br>What didn't you like? |
| How confident are you that you got to the right diagnosis or solution? | Was GPT better for diagnostic or management decision making, or equal? Why? |
| How efficiently do you feel you got to the diagnosis/solution you did? | Did GPT slow down or speed up your clinical decision making? |
| In what ways did GPT change your usual process for clinical decision making? | What about working with Chat GPT was useful?<br>Did working with Chat GPT feel like consulting a dictionary, talking to a colleague, or something else entirely?<br>Did it prompt you to take a different approach to clinical decision making?<br>Are there instances where ChatGPT surprised you with a unique perspective or solution to a diagnostic problem? |
| How trustworthy do you feel the outputs of GPT were? | In what ways could the trustworthiness of it be improved?<br>Were there specific situations where you disagreed with the AI's suggestions?<br>Did your trust in ChatGPT's suggestions evolve over the course of the study? How/Why? |
| **Integration into routine use**: My last set of questions are about how GPT or other AI tools might be integrated into routine use for clinical decision making. | |
| What do you think it would take to integrate AI into routine clinical decision making? | What things might help facilitate its use?<br>What do you see as the barriers to use? |

| | |
|---|---|
| Is there anything that concerns you about integrating Chat GPT or similar AI into routine clinical decision making? | E.g. ethics, patient perceptions, governance. |
| When there is a new innovation in clinical practice, do you like to be the first person to try it out, or do you prefer to adopt it later? (Innovator, early adopter, early majority, late majority, laggards) | |
| Is there anything else you'd like to add about the use of AI for clinical decision making that I haven't already asked about? | |
